# The HeartMagic prospective observational study protocol – characterizing subtypes of heart failure with preserved ejection fraction

**DOI:** 10.1101/2025.04.10.25325567

**Authors:** Philippe Meyer, Angela Rocca, Jaume Banus, Augustin C. Ogier, Costa Georgantas, Pauline Calarnou, Anam Fatima, Jean-Paul Vallée, Jean-François Deux, Aurélien Thomas, Julien Marquis, Pierre Monney, Henri Lu, Jean-Baptiste Ledoux, Cloé Tillier, Lindsey A Crowe, Tamila Abdurashidova, Jonas Richiardi, Roger Hullin, Ruud B. van Heeswijk

**Affiliations:** Cardiology Division, University Hospital of Geneva, Geneva, Switzerland; Cardiology Division, Lausanne University Hospital (CHUV), Lausanne, Switzerland; Department of Radiology, Lausanne University Hospital (CHUV) and University of Lausanne (UNIL), Lausanne, Switzerland; Radiology Division, Diagnostic Department, Geneva University Hospitals and University of Geneva, Geneva, Switzerland; Faculty Unit of Toxicology, CURML, Faculty of Biology and Medicine, University of Lausanne, Lausanne, Switzerland; Unit of Forensic Toxicology and Chemistry, CURML, Lausanne and Geneva University Hospitals, Lausanne, Geneva, Switzerland; Lausanne Genomic Technologies Facility, University of Lausanne, Lausanne, Switzerland; CIBM Center for Biomedical Imaging, Lausanne and Geneva, Switzerland

**Keywords:** Heart failure, HFpEF, CMR, quantitative MRI, genomics, machine learning, metabolomics, subtyping, echocardiography

## Abstract

**Introduction:** Heart failure (HF) is a life-threatening syndrome with significant morbidity and mortality. While evidence-based drug treatments have effectively reduced morbidity and mortality in HF with reduced ejection fraction (HFrEF), few therapies have been demonstrated to improve outcomes in HF with preserved ejection fraction (HFpEF). The multifaceted clinical presentation is one of the main reasons why the current understanding of HFpEF remains limited. This may be caused by the existence of several HFpEF disease subtypes that each need different treatments. There is therefore an unmet need for a holistic approach that combines comprehensive imaging with metabolomic, transcriptomic and genomic mapping to subtype HFpEF patients. This protocol details the approach employed in the HeartMagic study to address this gap in understanding.

**Methods:** This prospective multi-center observational cohort study will include 500 consecutive patients with actual or recent hospitalization for treatment of HFpEF at two Swiss university hospitals, along with 50 age-matched HFrEF patients and 50 age-matched healthy controls. Diagnosis of heart failure is based on clinical signs and symptoms and subgrouping HF patients is based on the left-ventricular ejection fraction. In addition to routine clinical workup, participants undergo genomic, transcriptomic, and metabolomic analyses, while the anatomy, composition, and function of the heart are quantified by comprehensive echocardiography and magnetic resonance imaging (MRI). Quantitative MRI is also applied to characterize the kidney. The primary outcome is a composite of one-year cardiovascular mortality or rehospitalization. Machine learning (ML) based multi-modal clustering will be employed to identify distinct HFpEF subtypes in the holistic data. The clinical importance of these subtypes shall be evaluated based on their association with the primary outcome. Statistical analysis will include group comparisons across modalities, survival analysis for the primary outcome, and integrative multi-modal clustering combining clinical, imaging, ECG, genomic, transcriptomic, and metabolomic data to identify and validate HFpEF subtypes.

**Discussion:** The integration of comprehensive MRI with extensive genomic and metabolomic profiling in this study will result in an unprecedented panoramic view of HFpEF and should enable us to distinguish functional subgroups of HFpEF patients. This approach has the potential to provide unprecedented insights on HFpEF disease and should provide a basis for personalized therapies. Beyond this, identifying HFpEF subtypes with specific molecular and structural characteristics could lead to new targeted pharmacological interventions, with the potential to improve patient outcomes.

## Introduction

### Background and Rationale

Heart failure (HF) is a major public health concern that leads to substantial morbidity, mortality, and healthcare costs, with a prevalence nearing 3% of the general population in most European countries.^1^ HF is classified into three categories based on left-ventricular ejection fraction (LVEF): “HF with reduced ejection fraction” (HFrEF) when LVEF is ≤40%, “HF with mildly reduced ejection fraction” (HFmrEF) when LVEF is between 41% and 49%, and “HF with preserved ejection fraction” (HFpEF) when LVEF is ≥50%.^2,3^ Over the past 30 years, HFpEF has become the most prevalent HF phenotype in Western countries, now accounting for over 50% of cases.^1,4^ The prognosis of patients hospitalized for HFpEF is poor, with reported 1-year all-cause mortality rates of ∼20-25% and rehospitalization rates approaching 50%.^1^

Many patients presenting with HF and an LVEF ≥50% exhibit specific cardiac conditions, such as infiltrative, hypertrophic, or restrictive cardiomyopathy, as well as valvular or pericardial diseases, which are now referred to as “HFpEF mimics”^5^ or “secondary HFpEF”.^6^ Diagnosis of HFpEF should therefore exclude these mimics. However, earlier HFpEF studies may have inadvertently included such patients,^7^ and even recent cohorts may still include misclassified cases due to incomplete investigation in elderly patients.

Even when mimics are excluded, HFpEF remains a heterogeneous syndrome.^6^ Traditionally, LV diastolic dysfunction has been considered central, with increased LV filling pressures and reduced cardiac output at rest or during exercise. More recently, systemic endothelial inflammation, driven by comorbidities such as hypertension, obesity, diabetes, COPD, sedentary lifestyle, or iron deficiency, has emerged as a unifying concept.^8–10^ This inflammation is linked to myocardial fibrosis, oxidative stress, and signaling pathway alterations that affect cardiomyocyte function.

Beyond diastolic dysfunction, other mechanisms contribute to HFpEF, including LV systolic dysfunction, impaired left atrial function, pulmonary vascular disease, right ventricular dysfunction, systemic arterial stiffening, coronary and peripheral microvascular dysfunction, chronotropic incompetence, skeletal muscle dysfunction, and arrhythmias, especially atrial fibrillation.^11,12^

Despite numerous large phase 3 clinical trials, most pharmacologic interventions have failed to demonstrate clear benefit in HFpEF.^13^ While sodium-glucose cotransporter-2 (SGLT2) inhibitors and the mineralocorticoid receptor antagonist finerenone have shown promise in HFmrEF and HFpEF,^14–16^ no trial has yet demonstrated a significant reduction in overall or cardiovascular mortality in HFpEF.^17^ This may be due to underlying pathophysiological heterogeneity,^6,18,19^ inclusion of HFpEF mimics,^7^ or the slow progression of the disease that requires longer study durations.^18^

Efforts by major societies, including the Heart Failure Association (HFA) of the European Society of Cardiology (ESC), have focused on defining phenotypes to guide targeted therapies.^6^ The HFA algorithm proposes seven HFpEF phenotypes with corresponding treatment strategies. Examples of phenotype-specific treatments include semaglutide (a Glucagon-Like Peptide-1’ (GLP-1) receptor agonist) and tirzepatide (a GLP-1 and glucose-dependent insulinotropic polypeptide (GIP) agonist), which have recently shown promise in treating the obese phenotype of HFpEF patients.^20–23^ However, substantial phenotype overlap limits the precision of such classifications. New strategies that extend beyond routine clinical data are therefore necessary to fully capture the complex heterogeneity of HFpEF disease.

### Objectives

This observational study, named HeartMagic (HEART failure studied with a MAchine learning, Genomics, and Imaging Combination), aims to perform comprehensive subtyping of HFpEF patients using a multimodal, integrative approach. By combining clinical data with genomics, metabolomics, and advanced cardiac magnetic resonance imaging (MRI), we will generate a rich, high-dimensional dataset.^13^ MRI enables detailed quantification of cardiac structure, function, hemodynamics, and tissue composition,^24^ while omics data provide insights into underlying molecular mechanisms. Together, these data form an ideal foundation for the application of machine learning (ML) models, which have shown transformative potential in healthcare by uncovering complex patterns and enhancing predictive accuracy.^25^

Our objective is therefore to identify clinically meaningful HFpEF subtypes linked to distinct pathophysiological mechanisms and outcomes. We aim to build one of the largest and most comprehensive HFpEF cohorts to date, supporting the development of more targeted and effective therapeutic strategies. This protocol outlines the methods used across all modalities.

**Central Figure.**
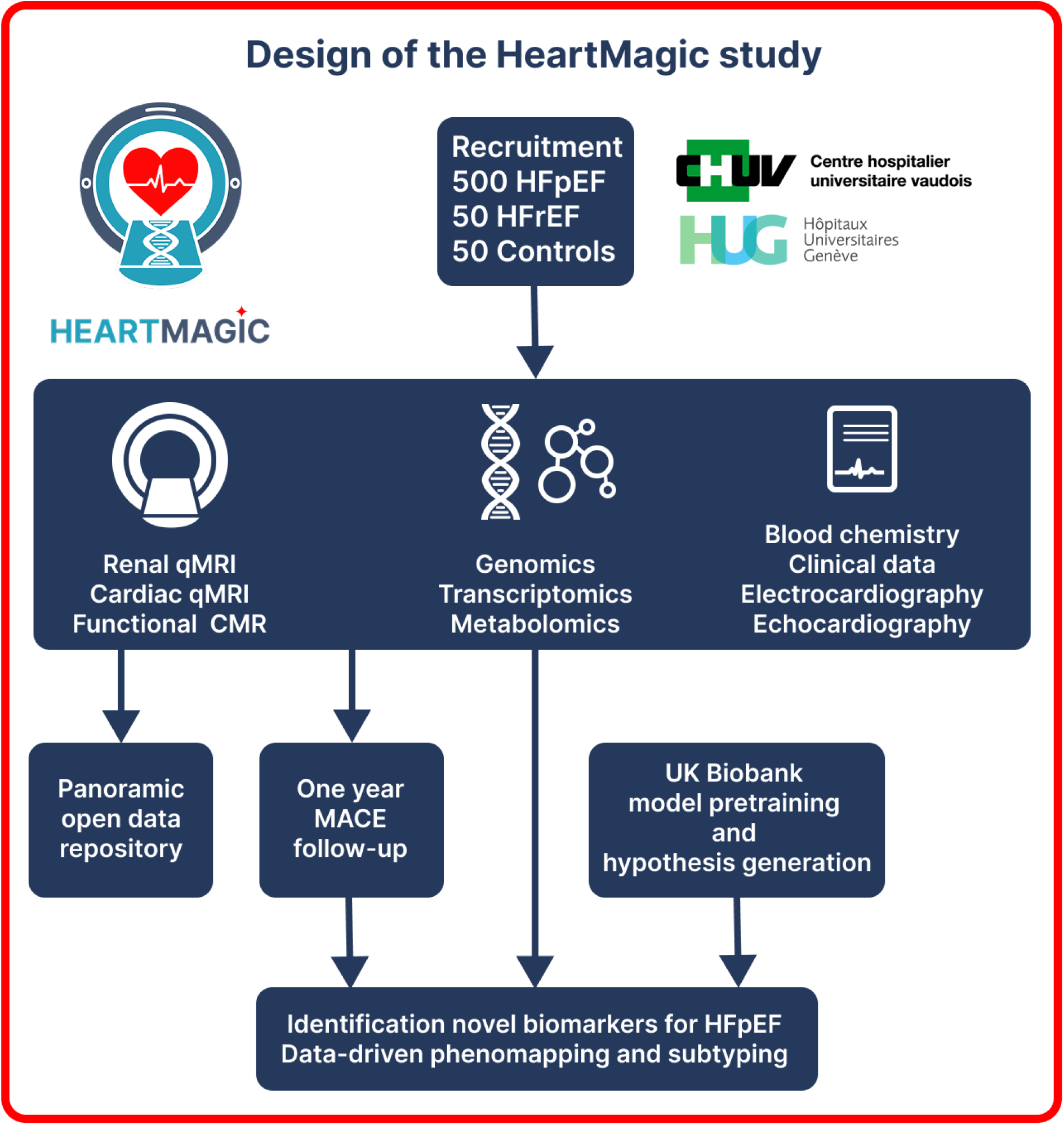
Setup and design of the HeartMagic study.

## Methods

### Study Design

This prospective observational clinical study will not interfere with standard care, and no randomization is required. A total of 500 consecutive patients with current or recent hospitalization for acute heart failure and preserved LVEF at the Geneva University Hospitals (HUG) or the Lausanne University Hospital (CHUV) will be enrolled, along with 50 HF patients with reduced LVEF and 50 age-matched healthy controls. The latter two groups will only serve as reference groups to enable comparison and calibration of multimodal analyses, while the primary aim of the study is to subtype HFpEF patients. These comparator groups will thus not be further stratified. Recruitment is planned over a 3-year period based on acute HF hospitalization rates at both centers. Each participant will undergo genetic, genomic, and metabolomic analyses, comprehensive echocardiography, and quantitative MRI. HF patients will receive a follow-up by phone at 30, 90 days and one-year post-discharge. Anonymized data will be made publicly available in persistent repositories when possible (see Open Access and Open Data further down).

### Setting

The study is conducted at two Swiss academic centers: the Geneva University Hospitals (HUG) and the Lausanne University Hospital (CHUV). HUG has approximately 2,000 beds, while CHUV has roughly 1,500 beds. Both are tertiary care facilities that serve a population of over 2 million people across their respective regions. Each year, approximately 1,200 patients are hospitalized for acute heart failure at HUG, while CHUV admits around 700 patients for the same condition according to the Swiss Federal Office of Statistics.

### Patients

Ethics approval was obtained from the Ethics Committee of the Canton of Vaud (CER-VD) of Switzerland under number 2022-00934. All participants provide written informed consent to participate prior to enrollment. The study will prospectively include 500 HFpEF patients, while simultaneously recruiting 50 HFrEF patients and 50 healthy participants, age- and sex-matched to the HFpEF group (Table 1). The latter two groups will serve as reference groups to enable comparison and calibration of multimodal analyses, while the primary aim of the study is to subtype HFpEF patients.

**Table 1.**
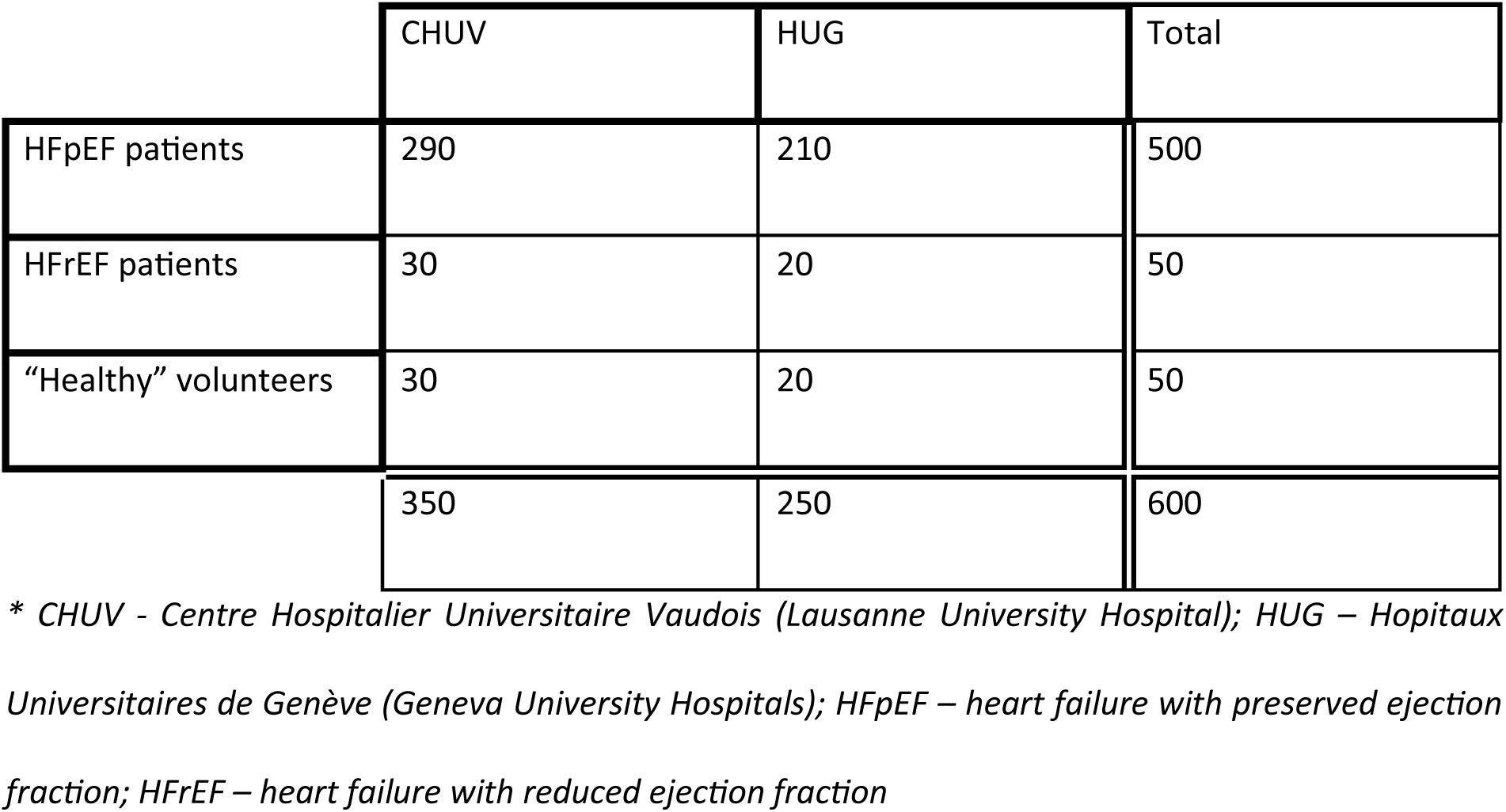
The expected distribution of recruitment across the two sites.

For patients with HFpEF the inclusion criteria are:

1. Adult ≥18 years old
2. Written and informed consent
3. Presence of dyspnea
4. Current or recent hospitalization for acute HF with signs of pulmonary and/or systemic congestion
5. On diuretic treatment prior to enrollment
6. Left ventricular ejection fraction (LVEF) ≥50% on echocardiography with at least 1 structural and/or functional cardiac abnormality compatible with HFpEF according to the current HFpEF definition.^3,26^

The exclusion criteria are:

1. Claustrophobia, ferromagnetic implants, or any other contra-indications to MRI
2. Contraindication to adenosine/regadenoson and/or gadolinium-based contrast agents
3. Pregnant or breastfeeding women
4. Alternative causes of dyspnea: valvular dysfunction considered by the investigators to be clinically significant, anemia with hemoglobin <10 g/dl, severe pulmonary disease
5. Chronic inflammatory disease of systemic or rheumatic origin
6. Cardiac amyloidosis or other infiltrative cardiac disease, sarcomeric hypertrophic cardiomyopathy, pericardial constriction
7. Occurrence of any of the following prior to enrollment:

- Acute inflammatory heart disease (e.g., myocarditis, endocarditis) within 90 days
- Myocardial infarction within 90 days
- Coronary artery bypass graft surgery within 90 days
- Percutaneous coronary intervention within 30 days
- Stroke or transient ischaemic attack within 90 days
8. Comorbid conditions associated with life expectancy <1year
9. Alcohol abuse
10. Liver cirrhosis classified as Child–Pugh C
11. Severe chronic kidney disease with eGFR <30 ml/min/1.73m2
12. Incapacity of judgement
13. Active Covid-19 infection
14. Unwillingness to receive notice of accidental findings
15. A body size prohibitive for the 60cm-diameter bore of the MR scanner

HFrEF patients are age- and gender-matched with HFpEF patients and share similar inclusion criteria, except for an LVEF <40%, as well as comparable exclusion criteria. Healthy volunteers (aged ≥60 years) must have NT-proBNP levels at or below the age-adjusted upper limit and no history of cardiac disease or more than moderate hepatic disease. They are also age- and gender-matched with the HFpEF group and share similar exclusion criteria with the other two groups.

### Recruitment

Patients hospitalized for dyspnea at the emergency department (ED) are screened for eligibility. The screening process includes an evaluation of heart failure symptoms and signs, underlying etiology, past medical history, initial blood test results, echocardiographic evaluation of LVEF, and the potential presence of cardiovascular implantable electronic devices and other ferromagnetic materials. Patients who meet the inclusion criteria are approached after their referral to the internal medicine and cardiology wards. They receive detailed information about the study and an informed consent form, with up to 48 hours allowed for consideration and decision-making.

The scheduled enrolment of 500 HFpEF patients, 50 HFrEF patients, and 50 healthy controls is expected to be complete by the end of 2026. All three participant groups will be recruited simultaneously. Weekly surveillance of age and sex distribution among recruited patients will be performed to document representativeness. Based on epidemiological data, we anticipate a female-to-male ratio of approximately 2:1 in HFpEF and 1:4 in HFrEF, but no predefined sex ratio will be enforced. For the HFrEF and healthy control groups, we will aim to maintain broadly comparable distributions to facilitate interpretability.

Age-matched healthy controls are recruited via the University for Seniors in Lausanne and Geneva through online advertisement. Interested individuals attend an informational session, complete the informed consent process, and receive a compensation of CHF 200.

### Patient Study Procedure

Each patient is invited to remain in follow-up for up to 1 year after inclusion, which aims to document routine care. The study procedures are divided into three parts (Figure 1):

1. Enrollment and Information visit: the patient receives information about the study’s purpose, procedures, risks, and benefits. After reviewing and discussing the informed consent form (ICF), the patient has the opportunity to ask questions before deciding to participate. If the patient agrees, they sign the ICF, and the next steps are arranged.
2. Baseline visit (Day 1): blood (∼40 mL) are collected. Transthoracic echocardiography and cardiac MRI are also performed on the same day.
3. Follow-up at Days 30, 90, and 365: Follow-up is conducted by phone and documents patient-related outcomes (morbidity, mortality, health status changes), which are validated through review of medical documentation in a second step. If a health issue has arisen, the participant is invited for an additional ambulatory visit at CHUV/HUG.

**Figure 1.**
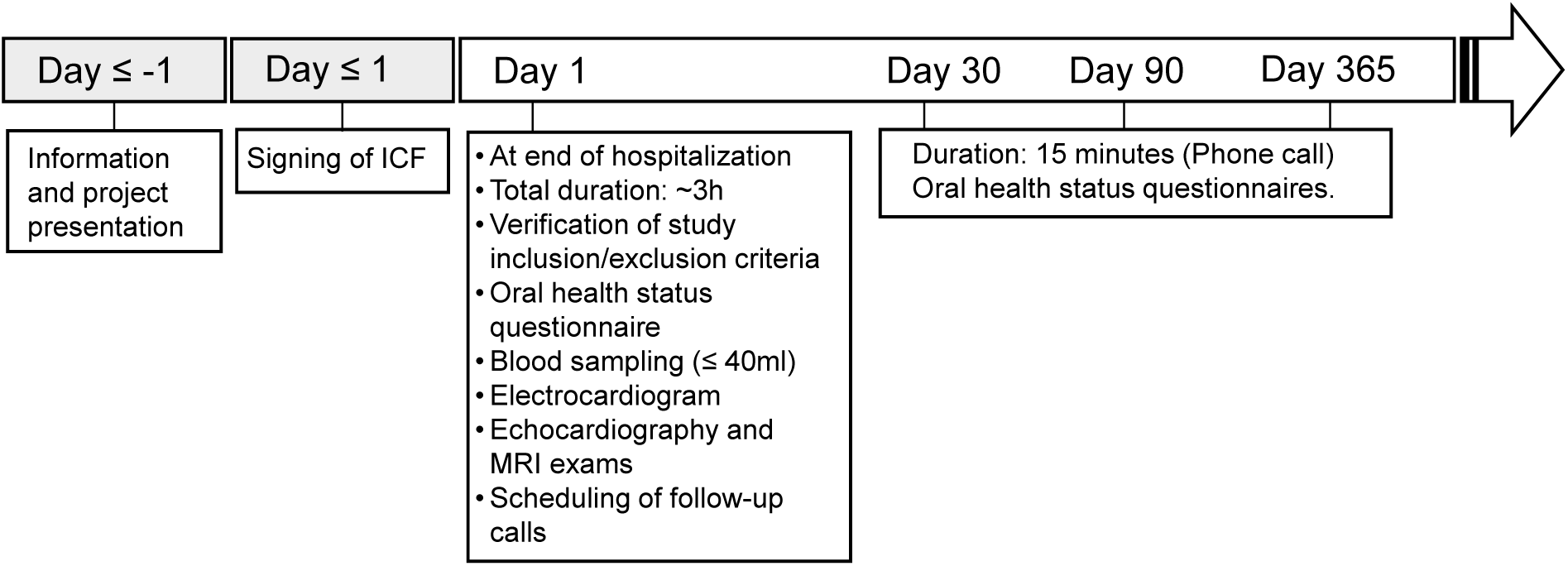
Flow chart of the study procedure. ICF – informed consent form; MRI – magnetic resonance imaging.

### Blood Sampling

Blood samples are collected for the analysis of metabolomics, genomics, transcriptomics, and clinical chemistry. After cubital venipuncture, specific tubes are filled for each type of analyses. Blood for clinical chemistry and hemogram analyses are processed by the local Departments of Laboratory Medicine (Supplementary Table 1). Samples for other analyses are aliquoted, frozen, and stored at - 80°C, and later sent in batches of 96 samples to specific laboratories. The assessment of blood parameters associated with systemic inflammation uses the human proinflammatory panel I (MESO SCALE DISCOVERY®) cytokine assay (K15052D), which measures interferon γ (IFNγ), interleukin-1ß (IL-1ß), interleukin-6 (IL-6), interleukin-10 (IL-10), and tumor necrosis factor α (TNFα).^27^ Additional aliquots of plasma and serum will be stored at –80 °C for potential future analyses, including measurement of novel biomarkers.

### Metabolomics

A total of 2.5 mL of whole blood is collected in lithium heparin tubes and centrifuged to obtain plasma, which is aliquoted into tubes suitable for −80°C storage. Samples are sent in batches of 96 to the University Center for Legal Medicine of Lausanne and Geneva (CURML) for analysis. Metabolites are extracted from 100 µL of plasma using a cold methanol-ethanol-water (2:2:1) solvent mixture. Samples are analyzed in full scan mode (positive and negative polarities) using ultra-high-performance liquid chromatography coupled with a Thermo Scientific high-resolution xploris 120 mass spectrometer (HRMS) in RP (Kinetex C18, in positive and negative modes). Separation is performed in gradient elution mode with mobile phases in positive mode (H2O; 0.1% formic acid and MeOH; 0.1% formic acid) and in negative mode (H2O; 5mM Ammonium acetate + 0.25mM Ammonium fluoride and MeOH). This will allow detection of several thousand metabolite features, as described previously.^28,29^ The expected overlap of compounds between the different approaches is used as an internal validation to assess the adequacy of our results. Raw UPLC-HRMS data are processed using Compound Discover 3.3 software for peak detection, chromatogram alignment, and isotope annotation. To provide the highest quality data for bioinformatics mining, quality controls (i.e., representative pool of samples) and internal standards are used to assess overtime-analytical drift (i.e., batch effects).^30^ To tackle this issue, we will use a method called systematic error removal using random forest (SERRF) and “DBnorm”, which includes algorithms for preprocessing and removing technical heterogeneity from the data via a statistical model in a two-stage procedure.^31^ The determination of the metabolomic signature has the potential to highlight discriminant metabolites significantly different between subtypes of the HFpEF umbrella.

Over the last few years, several large-scale metabolomics studies on thousands of biological samples have been performed on the platform described above.^32,33^ The human metabolome has not been completely covered, nonetheless, the discriminant metabolites will be characterized using both their MSn fragmentation pattern and open-access libraries.^34^ With accurate mass measurement, a home-made script will first be used to identify discriminant metabolites according to a predefined m/z list of metabolites. This list, including around five thousand present and/or detected metabolites in biofluids, is built from the human metabolome database (HMDB) and updated from our own library. We additionally use HMBD (www.hmdb.ca), lipidmaps (www.lipidmaps.org), metlin (http://metlin.scripps.edu/index.php), and mzcloud (www.mzcloud.org), totaling over 40,000 entries. Metabolite identification will also be then performed using SIRIUS 4.7.4, a software that determines the most likely elemental composition of metabolites through the analysis of isotopic patterns and MS/MS fragmentation spectra, and CANOPUS, a computational tool for systematic annotation of compound chemical classes.^35,36^

### Genomics

A total of 1.2 mL of whole blood is collected in EDTA-K3E tubes, aliquoted into tubes suitable for −80°C storage, and sent in batches of 96 to INRAE (the French National Research Institute for Agriculture, Food, and Environment; Paris, France) for DNA extraction and genotyping. Each batch includes a mix of samples from HFpEF patients, HFrEF patients, and controls Genotyping is performed on the GENTYANE INRAE Clermont platform (UMR GDEC n°1095 INRAE/UCA, https://gentyane.clermont.inrae.fr/) using the Axiom Precision Medicine Diversity Array (PMDA) (Thermo Fisher Scientific, Waltham, MA, USA). This array provides over 93% marker overlap with the UK Biobank Axiom array in the categories of interest for polygenic risk scores (see below), and over 800K markers in the imputation grid, ensuring that we can match with UKB genomic data. Data are provided in CEL format.

### Transcriptomics

For gene expression, 2.5 mL of whole blood is stored in PAXgene tubes at −80°C, and sent to QiaGen (Hilden, Germany) on dry ice for RNA extraction. Sequencing is performed at the Lausanne Genomic Technologies Facility, University of Lausanne, Switzerland (https://www.unil.ch/gtf). Sequencing libraries are prepared from 500 ng of total RNA using the Illumina Stranded mRNA kit from Illumina. Globin depletion is performed using the FastSelect kit from Qiagen. Sequencing is carried out on an Aviti sequencer (Element Biosciences, San Diego, CA, USA) using single-end sequencing with 150 cycles, at a coverage of 25M reads per sample. Sequencing data is demultiplexed using the Bases2Fastq 2.0 software, quality checked using Fastqc v0.11.9 (https://www.bioinformatics.babraham.ac.uk/projects/fastqc/) and Fastq Screen,^37^ and finally provided in FASTQ format.

### Electrocardiography

Electrocardiograms (ECGs) are digitally stored and analyzed using the validated BRAVO and GLASGOW algorithms,^38–40^ enabling extraction of more than 300 quantitative features for detailed electrical phenotyping. These high-dimensional data will be integrated with imaging and genomic datasets for advanced analyses, including deep learning–based approaches and genome-wide association studies (GWAS). In addition, the Transformer-based polygenic risk score (PRS) approach will be explored to model complex genotype–phenotype relationships. In this PRS approach, we propose to use an 8-layer pre-norm transformer architecture to modify SNP effect estimates from linear methods and from a covariate model. After thresholding out low-effect size SNPs, they will be combined into blocks of size 2048, mapped, and then concatenated with covariates before being fed to the transformer. After decoding to recover the original dimensions, the modified effect sizes will be summed to form a single scalar, added to the output of the covariate model, and the loss with respect to the ground truth continuous phenotype will be computed with a smooth L1 norm.^41^

### Transthoracic Echocardiography

TTE is routinely performed once patients are euvolemic and comfortable in the supine position. All exams are conducted on an Epiq CVx-3D Ultrasound system (Philips Healthcare, Eindhoven, the Netherlands) using an X5-1c 3D transthoracic probe. A comprehensive 2D examination is performed initially.^42^ Additionally, 3D acquisitions of the left ventricle and left atrium are obtained using the Dynamic Heart Model automated 3D tool (Philips Healthcare, Eindhoven, the Netherlands), and 3D images of the right ventricle are captured using the 3D-autoRV tool (Philips Healthcare, Eindhoven, the Netherlands). Longitudinal strain of the left ventricle, left atrium, and right ventricular free wall is assessed using the Autostrain LV, Autostrain LA, and Autostrain RV postprocessing tools, respectively (Philips Healthcare, Eindhoven, the Netherlands). All measurements adhere to the recommendations from the European Association of Cardiovascular Imaging for chamber quantification,^43^ evaluation of left ventricular diastolic function,^44^ and the European consensus on the diagnosis and imaging of HFpEF.^26,45^ Valvular disease is classified as mild, moderate, or severe according to current guidelines.^46–48^ The list of echocardiographic measurements is presented in Supplementary Table 2.

### Magnetic Resonance Imaging

#### Image Acquisition

Study participants are scheduled for a one-hour MRI exam on a fully CE-marked clinical 3 tesla (T) scanner (Prisma or PrismaFit, Siemens Healthineers, Erlangen, Germany) of the Center for BioMedical Imaging (CIBM) located in the Departments of Radiology of the CHUV and the HUG. Scanning is always performed by a technologist trained for cardiac research scans or a radiologist specialized in cardiac MRI, and a cardiologist is present for the injection of the pharmacological stressor. Besides routine localizer scans, this one-hour exam subsequently consists of (**Table 2**):

1. Kidney morphology: Coronal and transverse T_2_ HASTE.
2. Kidney oxygenation: Breath-held coronal BOLD T_2_* mapping.
3. Kidney tissue characterization: free-breathing coronal native joint T_1_-T_2_ mapping (PARMANav – PArametric Radial MApping with respiratory NAVigation) to quantify interstitial fibrosis and edema.
4. Cardiac function: routine 2D cine of the heart (short-axis stack, two-chamber, three-chamber, four-chamber, and left-ventricular outflow tract views).
5. Cardiac flow: 2D flow just above the level of the aortic valve to measure the ejected blood volume.
6. Myocardial tissue characterization: free-breathing 2D native joint T_1_-T_2_ mapping (PARMANav) to quantify interstitial fibrosis and edema at the basal, mid-ventricular, and apical short-axis as well as the four-chamber orientations.
7. Stress myocardial perfusion: if eGFR>30mL/min/1.73m^2^, quantitative myocardial perfusion imaging (qPerf) after the injection of a pharmacological stressor (100 mg adenosine or 400 µg regadenoson) Either stressor is applied since they are part of the local protocols; in addition, their effect on the physiologically in equivalent.^49^ This is followed by a half-dose of gadolinium-based contrast agent (GBCA): either 0.05 mmol/kg gadobutrol (Gadovist, Bayer AG) or 0.1 mmol/kg gadoteric acid (Dotarem, Guerbet) during the acquisition.
8. Cardiac anatomy and function: A second half-dose of GBCA is immediately followed by free-running 5D imaging (5D FRF) to assess anatomy and function at isotropic high spatial resolution in addition to the routine 2D cine.
9. Thoracic flow: 4D (i.e. 3 spatial dimensions over time) flow imaging to quantify the flow patterns and velocities in the heart and great vessels.
10. Post-contrast myocardial tissue characterization: repetition of the free-breathing joint T_1_-T_2_ mapping 8-18min after the injection of the second dose of GBCA, now to quantify the extracellular volume (ECV).
11. Quantitative myocardial rest perfusion imaging: the same scan as the stress perfusion imaging, without the stressor and with a third half-dose of GBCA.

**Table 2.** MRI pulse sequence parameters.

| Sequence | HASTE | BOLD | PARMANav | 2D cine | qPerf | 5D FRF | 4D flow |
| --- | --- | --- | --- | --- | --- | --- | --- |
| Sequence type | TSE | ME-GRE | GRE | bSSFP | GRE | GRE | GRE |
| Dimensions | 2D | 2D | 2D | 2D | 2D | 5D | 4D |
| TE (ms) | 96 | 6-52 | 1.5 | 1.28 | 0.98 | 1.33 | 2.25 |
| TR (ms) | 1600 | 76 | 3.36 | 3.39 | 1.99 | 2.99 | 4.27 |
| Echoes | 205 | 12 | 1 | 1 | 1 | 1 | 1 |
| Flip angle (°) | 80 | 30 | 12 | 43 | 14 | 15 | 12 |
| Matrix size | 256x256 | 256x256 | 192x192 | 256x170 | 192x114 | 160x160<br>x160 | 160x102x<br>39 |
| Field of view<br>(mm<sup>2</sup>) | 280x280 | 400x400 | 300x300 | 380x285 | 360x270 | 220x220<br>x220 | 380x304x<br>120 |
| Slices | 10 | 1 | 1 | 1-12 | 3 | - | - |
| Slice thickness (mm) | 5 | 5 | 8 | 8 | 8 | 1.38 | 2.5 |
| Acceleration | GRAPPA, PF | - | CS | CS | GRAPPA, PF | CS | GRAPPA/CS |
| ECG triggering | N | N | Y | Y | Y | N | Y |
| Resp MoCo | BH | BH | LL-NAV | BH | FB | SN | LL-NAV |
| Temp resolution (ms) | - | - | 151 | HB/25 | 227ms | HB/25 | HB/25 |
| k-Space trajectory | Cart | Cart | GA-Rad | Cart | Cart | GA-Rad | Cart |
| Inline recon | Y | Y | N | Y | Y | N | Y |
| Acquisition duration | 16s | 23s | 25-40HB | 2-24s | 90HB | 4m05s | 4m37s-9m |
| Prep module(s) | - |  | T<sub>2</sub>p, IR | - | Sat | - | - |
| Reference | 52 | 53 | 54 | 55 | 56 | 24 | 57 |

\* HASTE – Half-Fourier-Acquired Single-shot Turbo Spin Echo, BOLD – blood oxygen level dependent, PARMANav – Parametric radial mapping with respiratory navigation, ME-GRE – multi-echo GRE, bSSFP – balanced steady-state free precession, BH – breath hold, GA – golden angle, LL-NAV – lung-liver respiratory navigator, FB – free breathing, HR – heart rate, HB – heartbeat, Cart – Cartesian, Rad – radial, IR – inversion recovery, T<sub>2</sub>p – T<sub>2</sub> preparation, GRAPPA – GeneRalized Autocalibrating Partial Parallel Acquisition, PF – partial Fourier, CS – compressed sensing

After the scan, images and raw k-space data are directly saved to secured servers managed by the CHUV and HUG information technology departments.

Late gadolinium enhancement (LGE) imaging for the visualization of myocardial scar tissue is not performed because it has been demonstrated that *small* focal scars do not affect the outcome of HFpEF,^50^ while larger scars are visualized on the post-GBCA T_1_ maps, ECV maps, and synthetic LGE maps.^51^

#### Image and Map Reconstruction

HASTE images and BOLD maps of the kidney as well as 2D cine, qPerf, and 4D flow of the heart are reconstructed on the scanner with manufacturer software. Raw data of the PARMANav and 5D FRF acquisitions are exported to a separate workstation for image and map reconstruction.

For the PARMANav joint T_1_-T_2_ maps of both the kidney and heart, source images are reconstructed with compressed sensing.^54,58^ The extended phase graph formalism is employed to simulate a dictionary of signals for a broad range of T_1_ and T_2_ values. This dictionary is acquisition-specific and must be generated for each scan, as heart rate and respiratory gating influence magnetization evolution. The reconstructed images are matched pixel-by-pixel to the closest dictionary entry using a dot product, resulting in T_1_ and T_2_ maps.

For the free-running 5D images, cardiac and respiratory motion signals are self-gated and extracted from the raw data using the repeated superior–inferior (SI) projections of the 3D radial trajectory.^59^ Principal component analysis is first applied to separate the respiratory components, and second-order blind identification is used to isolate the cardiac components.^60^ A Gaussian fit is applied to the power spectral density of each resulting component to identify the cardiac signals,^61^ which favors the extraction of signals with a fundamental frequency within the expected physiological range. These physiological signals are subsequently used to bin the acquired data into cardiac and respiratory phases as previously described,^24^ resulting in undersampled 5D data. Respiratory binning is achieved by dividing the respiratory signal into four equally populated phases, ranging from end-expiration to end-inspiration. The cardiac signal is used to organize the data into non-overlapping phases of 50 ms. Finally, a k-t sparse SENSE compressed sensing algorithm is employed to reconstruct the 5D whole-heart images.^24,62^ Cardiac and respiratory regularization weights are set to 0.005 and 0.03, respectively, to ensure optimal reconstruction quality while maintaining the temporal and respiratory coherence of the data.

### Clinical Data

We collect data related to hospitalization duration (timing), movements between the services, interventions, medication, and imaging exams from the hospital information systems (Supplementary Table 4). All clinical data are coded. Participant data from both hospitals are entered into an electronic case report form (eCRF). The study database is RedCap®, hosted on a secure institutional server at CHUV, with protected access.

### Data Analysis and Statistics

#### Overall strategy

The statistical analysis plan focuses on developing imaging and genomics analysis methods applicable to the UK Biobank (UKB), which is our discovery cohort, and applying these methods on our own data to validate our findings. While UKB is a community sample and not clinically enriched in HFpEF patients, its large size makes it suitable as a development and discovery dataset around heart failure, cardiovascular disease more generally, and HFpEF risk factors. Using the validated H2FPEF risk score as a surrogate for HFpEF diagnosis^63^ will be considered, although it is limited by several factors: 1) transthoracic echocardiography is not available in UKB, leading to a truncated maximum score. 2) the score is meant to be applied to patients presenting with dyspnea, rather than a generic population. Diagnostic uncertainty is further increased by lack of NT-proBNP values for UKB subjects.

Initially, we will use group comparisons to identify differences in each of the modalities between the different clinical groups available in our dataset, as well as unsupervised methods within the HFpEF group. In a second step, we will resort to survival analysis (time-to-event analysis for the primary outcomes of the study), in particular within the HFpEF group, to focus on clinically relevant disease subtypes. Finally, we will focus on integrative analysis, where multiple modalities (clinical, imaging, ECG, genomics, transcriptomics, metabolomics…) are considered jointly for subtyping.

#### MR Image and Map Analysis

Renal diameters and volume, medullary and cortical thickness and volume are derived from the HASTE series. In the renal T_1_, T_2_, and T_2_* maps the cortex and medulla are automatically segmented with a deep-learning-based 2D U-Net.^64^

In the routine 2D cine images, LV and RV blood and myocardial volumes and mass, LVEF, RVEF, LA and RA volumes are manually segmented. The results in the first 100 subjects are then used to train DL-based automated segmentation networks for the 2D cines. LV long-axis systolic function is measured using mitral annular systolic excursion and LV global longitudinal strain (GLS). Left and right total atrial emptying fraction, passive atrial emptying fraction and active atrial emptying fraction are measured from the maximum LA and RA volumes, the prior to atria contraction and the minimum atrial volumes as reported by Kowallick et al.^65^

We also use these 2D cine data to develop methods to study spatio-temporal patterns. We process the images, segment the ventricles and divide the myocardium in different regions (16 AHA sub-segments for the left ventricle myocardium, 9 segments for the right ventricle myocardium). These divisions give us the flexibility to represent the heart as a spatio-temporal graph in which the nodes represent these myocardium regions and edges represent similarities between segments. Used across multiple acquisition sequences (relaxometry, strain, motion, etc.), these structural heart graphs will represent quantitative tissue properties, mechanical properties, and structural properties of the heart using a unified representation. The goal is to identify key functional and anatomical properties that are relevant to quantify the cardiac function and the cardiovascular health. The use of graph neural networks will allow the use of low-dimensional, latent representations of structure and function.

Similarly, from the 5D FRF images^24^ we extract global measures such as the diastolic and systolic atrial and ventricular volumes, as well as segmental myocardial measures such as wall thickness and strain. To assess the presence of pulmonary hypertension for the H2FPEF composite score,^63^ we calculate the septal-to-free-wall curvature ratio.^66^

Myocardial T_1_ and T_2_ values are manually segmented on the PARMANav maps according to the standardized 17 AHA LV segments,^67^ as well as for their equivalent RV segments where available.^68^ Subendocardial and subepicardial as well transmural myocardial perfusion is quantified in 16 sectors at 3 level (basal, mid and apical) according to the Kellman method.^69^

In the 4D flow MRI we measure aortic and pulmonary flow, flow through the atrioventricular and ventriculo-arterial valves, the transmitral flow profile, wall shear stress (WSS) at the level of the aorta and pulmonary artery, and pulse wave velocity (PWV) in the aorta. An analysis of the components of intraventricular flow and turbulence in the left atrium are performed on a dedicated software (Cvi 42, Circle, Montreal, Canada). Peak early diastolic tissue velocity (e’) is assessed at the septal and lateral mitral annulus using automated mitral valve tracking. Mitral inflow velocities (E and A) are measured and combined with e’ to measure the average septal-lateral E/e’ ratio reflecting pulmonary capillary wedge pressure. Tricuspid annular plane systolic excursion (TAPSE) and peak tricuspid annular systolic tissue velocity (S’) is measured as indices of right ventricular (RV) longitudinal systolic function.

To identify associations between the image-based biomarkers and clinical outcomes we will further develop systematic screening approaches with careful propensity matching and survival analysis^70^ to relate these potential biomarkers to clinical outcomes.

#### Omics Preprocessing and Analysis

For genomic data, we will apply standard quality control (QC) and preprocessing^71^ including sample quality checks (sex inconsistencies, call rate…), marker quality checks (minor allele frequency, Hardy-Weinberg equilibrium), and batch effect analysis and correction, using the bigsnpr package^72^ and other in-house tools.

For metabolomic data, after acquisition, preprocessing and QC steps described above, we will use the negative and positive mode results separately (around 8000 metabolites in total), obtained from untargeted analysis, to narrow down the list to around 200 metabolites for in-depth characterization.

This will be done by clinical group comparison, as well as correlation screening with respect to imaging features of interest, survival-linked features of interest, and associations with selected genes and SNPs.

For transcriptomic data, we will run an established QC pipeline (RNA-SeQC2^73^), annotate to the latest genome assembly, and quantify gene expression using Salmon^74^ (using the alternative pipeline of first aligning with STAR^75^ and quantifying using HTSeq)^76^.

#### Novel Biomarkers for HFpEF

Each omics will be processed according to modality-specific pipelines as described above. For multimodal analysis, our first approach will be to group participants according to one modality (for example, clustering by above- vs below-median CMR-derived markers and studying related survival), then examine the distribution of other biomarkers between these groups (for example, metabolomics profiles). To promote ‘early integration’ of modalities, in our second approach we will construct participant similarity graphs for each modality, where each participant is a node and edges represent pairwise similarity (e.g., cosine distance between expression values of candidate genes). Each such similarity graph will be treated as a “layer” in a multilayer graph, and multimodal subtypes will then be defined through community detection algorithms such as the Leiden algorithm. As a baseline benchmark for these approaches, we will use Similarity Network Fusion. The resulting multimodal communities will subsequently be evaluated by survival analysis.

The main focus will be to find biomarkers with outcomes-linked differences within the HFpEF group, in particular using survival analysis (“survival subtypes”). Because the follow-up period is one year and the recruitment is ongoing, during the time period where event counts are insufficient for modelling, we will in the initial analysis phase focus instead on unsupervised subtyping (“phenotyping subtypes”) and supervised group characterization. Then, when outcomes are observed, clustering results will be evaluated by looking at the silhouette coefficient, the concordance index, normalized mutual information (NMI), and the adjusted Rand index (ARI) with respect to observed primary and secondary clinical outcomes.

Another approach to mitigate the lack of events is to rely on the UK Biobank (UKB), a large observational cohort, for hypothesis generation. We use the UKB dataset to identify various CMR-derived biomarkers that have prognostic value on the risk of hospitalization and death due to heart failure, as well as for co-morbidities associated with HFpEF. These analyses will serve to narrow down the set of candidate biomarkers to a manageable number that can be tested with sufficient power in the HeartMagic cohort. For genomics specifically, we will derive low-dimensional risk scores from GWAS in UKB and validate them in HeartMagic, thereby ensuring feasibility despite smaller sample sizes.

A focus of the work is imaging genetics– using CMR-derived imaging phenotype as complex traits and GWAS targets. Preprocessed genomics data will thus be used for GWAS analyses (plink tool), in UKB for discovery, and our sample for validation. While cine and T_1_ acquisitions are not the same in both datasets, the approach should allow us to narrow down the set of plausible candidates. From these, we will also generate low-dimensional representations of genomic data, to be included in subtyping analyses, by using our newly proposed transformer-based non-linear method for polygenic risk scoring.^77^

Throughout, we will use a combination of the omics available to obtain a better understanding of group differences, phenotyping subtypes, and survival subtypes. For example, we will look at consensus across omics, and look at e.g. differential gene expression between subtypes obtained from other omics and modalities, although this approach can struggle to exploit discordant relationships^78^. We will also pursue graph-based approaches, viewing the problem as a community detection problem on a multilayer graph.

These analyses aim to produce novel interpretable, predictive and reproducible biomarkers that can improve clinical decision making, refine prognostic estimates, and can be used to propose novel treatment avenues and secondary endpoints for future clinical trials.

### Open Access and Open Data

All studies and sub-studies will be published in an open access format. All data will be converted to depersonalised non-proprietary formats and will be deposited in FAIR online repositories for open access, except for genomic data, which will be deposited with access control.

Identifying attributes including names and dates of birth will be removed from all data. Then, data will be depersonalized according to its type: tabular data (clinical, blood chemistry, ECG features) will be depersonalized using tools such as ARX;^79^ imaging data will be converted to Nifti before sharing, with potentially sensitive headers removed; sequence reads in transcriptomic data will be sanitized;^80,81^ metabolomic data will be shared as is since is not thought to contain directly identifying information.

## Discussion

This study protocol addresses the complex and heterogeneous nature of HFpEF, a prevalent condition with poor prognosis and few effective pharmacological treatments. By integrating advanced MRI with metabolomic, transcriptomic, and genomic analyses, this prospective observational study aims to refine HFpEF subtyping through machine learning (ML)-based multimodal clustering. With a cohort of 500 HFpEF patients, 50 age-matched HFrEF patients, and 50 healthy controls, we seek to identify clinically meaningful subgroups linked to distinct pathophysiological mechanisms and clinical outcomes, with a primary composite endpoint of one-year cardiovascular mortality or rehospitalization.

ML is central to precision medicine, particularly when applied to rich panoramic data such as genetics, metabolomics, proteomics, and quantitative MRI.^78^ This study focuses on two ML strategies: integrative representations,^82^ which merge omics and imaging data for improved disease characterization, and multimodal subtyping, which enhances phenotypic resolution. Graph-based representations, known for their interpretability and expressiveness, will play a key role in advancing core ML methodologies. Notably, network analysis has been described as a “grand unifier” in biomedical data science, underscoring its potential in HFpEF research.^83^

Identifying clinically relevant HFpEF subtypes remains a challenge, as prior subgroup analyses based on conventional parameters have yet to yield effective therapeutic targets. Evidence from cancer research suggests that integrating multiple biological measurements before subtyping can yield more consistent and clinically actionable classifications.^84^ By applying this principle to HFpEF, we aim to overcome previous limitations and improve phenotyping strategies.

The inclusion of a large, well-characterized cohort, along with comparative groups of HFrEF patients and healthy controls, strengthens the study’s generalizability and analytical depth. Furthermore, its prospective design and clinically relevant primary outcome measure ensure robust validation of the identified HFpEF subtypes, with the ultimate goal of advancing personalized treatment strategies.

Beyond the HFpEF pathology, it is expected that the integrative methods developed during this project will translate to other cardiovascular conditions and will also be relevant to several MRI radiogenomics studies, such as those involving hepatocellular carcinoma.

### Feasibility

The study protocol for this HFpEF project is highly feasible, supported by the multidisciplinary expertise of four main investigators and several project partners, each contributing complementary skills to manage the protocol’s complexity. Ruud van Heeswijk leads a radiology-embedded engineering team at CHUV, ensuring access to advanced MRI technology, secure data storage, and computational resources. Jonas Richiardi heads the translational machine learning lab, specializing in medical imaging techniques. Roger Hullin and Philippe Meyer lead the Heart Failure Units at CHUV and HUG, respectively, overseeing nearly 1,000 HFpEF patient cases each year. Each investigator directs one of three Work Packages: van Heeswijk (MRI), Richiardi (machine learning), and Hullin and Meyer (clinical study). Defined roles and collaborative structures across teams ensure effective data handling, patient recruitment, and ethical compliance, with research nurses in Hullin’s and Meyer’s units managing recruitment and follow-up. In addition, project partners provide required expertise, for example in statistical genetics (Zoltan Kutalik and team), metabolomics (Aurélien Thomas and team), transcriptomics (Julien Marquis and team), advanced cardiac MRI (Matthias Stuber and team), clinical thoracic and abdominal imaging (Jean-Paul Vallée and team), and 4D flow (Jean-François Deux). The project is also supported by the Clinical Trial Unit at CHUV, ensuring methodological rigor and regulatory compliance. This well-integrated approach provides the necessary capabilities to address the complexities of HFpEF phenotyping and ensures the feasibility of recruitment, data acquisition, and analysis.

### Strengths and Limitation

As an observational study, our work is limited by its inability to establish causal relationships between identified HFpEF subtypes and outcomes. Additionally, for resource and logistical reasons, follow-up is limited to one year and potential medium- and longer-term outcomes will not be observed.

Our inclusion and exclusion criteria may limit the representativity of the sample, in particular with respect to comorbidities, which may be more prevalent in the general HFpEF population than in our sample. Likewise, while the study is already multi-centric, its focus on western Switzerland limits the representativity of our sample on a wider geographical scale. However, Switzerland is one of the most multicultural countries in Europe, with an estimated 40% of the population having a migration background (either foreign nationals or Swiss-born individuals with foreign-born parents). While the control and HFrEF groups are relatively small, they are not intended for detailed subgroup analyses but rather as comparator cohorts. The primary discovery aim of the study lies within the HFpEF group. With respect to outcomes, assuming a 12-month cardiovascular mortality rate of 15% and a re-hospitalization rate of 50% within HFpEF, we anticipate approximately 75 deaths and 250 re-hospitalizations. Models will be evaluated both in UK Biobank and in HeartMagic, using cross-validation and independent training/testing schemes. Even under conservative assumptions (e.g. sensitivity of 70% for mortality classification), the lower bound of the 95% confidence interval remains well above chance, supporting the adequacy of the planned sample size for biomarker discovery and validation.

A further limitation is that the analytical methods developed using UK Biobank (UKB) data may not directly transfer to the HeartMagic cohort, given the differences in study populations, imaging modalities, and healthcare systems. Nevertheless, evidence from cardiovascular imaging and other application domains^85^ shows that leveraging external datasets for hypothesis generation and model development can improve performance and robustness in smaller, disease-specific cohorts. UKB imaging data are already multicentric, which facilitates the learning of machine-independent features and supports generalization across settings.

The reliance on advanced imaging and molecular techniques may limit reproducibility in settings without similar resources, and the complexity of data analysis may pose challenges for real-time clinical application.

Despite these limitations, the study’s extensive dataset could serve as a foundational resource for future HFpEF research, with potential to guide targeted therapeutic developments.

**Table 3.**
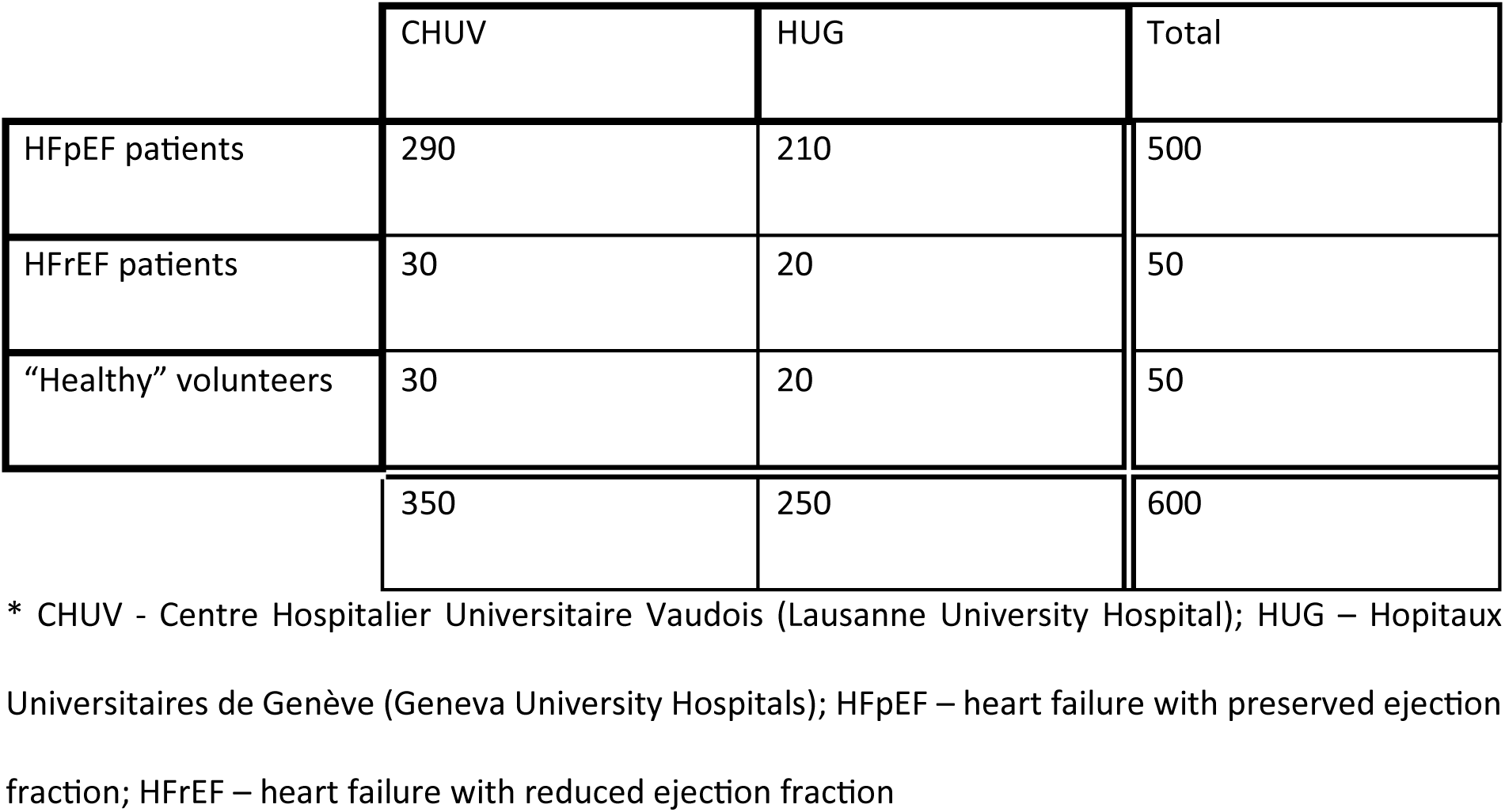
Expected distribution of recruitment across the two sites.

**Table 4.** MRI pulse sequence parameters.

| Sequence | HASTE | BOLD | PARMAN<br>av | 2D cine | qPerf | 5D FRF | 4D flow |
| --- | --- | --- | --- | --- | --- | --- | --- |
| Sequence type | TSE | ME-GRE | GRE | bSSFP | GRE | GRE | GRE |
| Dimensions | 2D | 2D | 2D | 2D | 2D | 5D | 4D |
| TE (ms) | 96 | 6-52 | 1.5 | 1.28 | 0.98 | 1.33 | 2.25 |
| TR (ms) | 1600 | 76 | 3.36 | 3.39 | 1.99 | 2.99 | 4.27 |
| Echoes | 205 | 12 | 1 | 1 | 1 | 1 | 1 |
| Flip angle (°) | 80 | 30 | 12 | 43 | 14 | 15 | 12 |
| Matrix size | 256x256 | 256x256 | 192x192 | 256x170 | 192x114 | 160x160x<br>160 | 160x102x<br>39 |
| Field of view<br>(mm<sup>2</sup>) | 280x280 | 400x400 | 300x300 | 380x285 | 360x270 | 220x220x<br>220 | 380x304x<br>120 |
| Slices | 10 | 1 | 1 | 1-12 | 3 | - | - |
| Slice thickness<br>(mm) | 5 | 5 | 8 | 8 | 8 | 1.38 | 2.5 |
| Acceleration | GRAPPA,<br>PF | - | CS | CS | GRAPPA,<br>PF | CS | GRAPPA/<br>CS |
| ECG triggering | N | N | Y | Y | Y | N | Y |
| Resp MoCo | BH | BH | LL-NAV | BH | FB | SN | LL-NAV |
| Temp<br>resolution (ms) | - | - | 151 | HB/25 | 227ms | HB/25 | HB/25 |
| k-Space<br>trajectory | Cart | Cart | GA-Rad | Cart | Cart | GA-Rad | Cart |
| Inline recon | Y | Y | N | Y | Y | N | Y |
| Acquisition duration | 16s | 23s | 25-40HB | 2-24s | 90HB | 4m05s | 4m37s-9m |
| Prep module(s) | - |  | T<sub>2</sub>p, IR | - | Sat | - | - |
| Reference | 52 | 53 | 54 | 55 | 56 | 24 | 57 |

\* HASTE – Half-Fourier-Acquired Single-shot Turbo Spin Echo, BOLD – blood oxygen level dependent, PARMANav – Parametric radial mapping with respiratory navigation, ME-GRE – multi-echo GRE, bSSFP – balanced steady-state free precession, BH – breath hold, GA – golden angle, LL-NAV – lung-liver respiratory navigator, FB – free breathing, HR – heart rate, HB – heartbeat, Cart – Cartesian, Rad – radial, IR – inversion recovery, T<sub>2</sub>p – T<sub>2</sub> preparation, GRAPPA – GeneRalized Autocalibrating Partial Parallel Acquisition, PF – partial Fourier, CS – compressed sensing

## Data Availability

This is a study protocol description; a Data Availability Statement does not apply.

## Acknowledgements

This research is conducted using the UK Biobank Resource under Application Number 80108.

## Funding Sources

The HeartMagic study is funded by the Swiss National Science Foundation (SNSF) under grant number CRSII5_202276.

## Conflict of Interest Disclosures

None.

## Supplemental Material

Supplementary Tables 1-4

## Abbreviations

CE: Conformité européenne
CHUV: Centre Hospitalier Universitaire Vaudois (Lausanne University Hospital)
CMR: Cardiovascular Magnetic Resonance
eCRF: Electronic Case Report Form
EF: Ejection Fraction
HEARTMAGIC: HEART failure studied with a MAchine learning, Genomics, and Imaging Combination
HFpEF: Heart Failure with Preserved Ejection Fraction
HFrEF: Heart Failure with Reduced Ejection Fraction
HUG: Hôpitaux Universitaires de Genève (Geneva University Hospitals)
ICF: Informed Consent Form
LV: Left Ventricle
ML: Machine Learning
NT-proBNP: N-terminal pro–B-type Natriuretic Peptide
PARMANav: PArametric Radial MApping with respiratory NAVigation
qMRI: Quantitative Magnetic Resonance Imaging
qPerf: Quantitative Perfusion
RV: Right Ventricle
UKB: UK Biobank

## Additional Declarations

### Ethics approval and consent to participate

Ethics approval was obtained from the Ethics Committee of the Canton of Vaud (CER-VD) of Switzerland under number 2022-00934. All participants provide written informed consent to participate prior to enrollment.

### Consent for publication

All authors consent to this publication.

### Availability of data and materials

It is planned to make all data available in a public repository after anonymization; this is detailed in the informed consent forms that all participants signed.

### Authors’ contributions

Meyer, Richiardi, Hullin, and van Heeswijk wrote the initial draft of the manuscript. All authors significantly revised the manuscript. Van Heeswijk coordinates the study. Meyer and Tillier set up the recruitment and screening in Geneva; Hullin, Richiardi, Rocca, and Abdurashidova set up the recruitment and screening in Lausanne. Rocca set up the coordination between the project partner teams. Rocca, Tilier and Hullin set up the blood sampling and storage. Thomas established the metabolomic analysis. Marquis, Hullin, and Richiardi set up the genotyping and transcriptomics protocol. Hullin established the ECG analysis. Meyer and Monney set up the echocardiography protocol. Van Heeswijk, Ledoux, Vallée, Ogier, Crowe, Calarnou, Fatima, and Deux set up the MRI acquisition, reconstruction, and analysis. Rocca, Tillier, Meyer, and Hullin collected the clinical data. Richiardi, Banus, and Georgantas established the multi-modal data integration and analysis procedure.

### Authors’ information (optional)

N/A

## Notes

### Competing Interest Statement

The authors have declared no competing interest.

### Funding Statement

The study is funded by the Swiss National Science Foundation (SNSF) under grant number CRSII5_202276.

### Author Declarations

The Ethics Committee of the Canton of Vaud (CER-VD) of Switzerland gave ethical approval for this work under number 2022-00934. All participants provide written informed consent to participate prior to enrollment.

### Summary of Updates

The manuscript was submitted to the Journal of the American Heart Association, where a Revision was requested. The updated version is the result of this revision.

