## Supplemental Tables 1-4 for "The HeartMagic prospective observational study protocol – characterizing subtypes of heart failure with preserved ejection fraction"

### Supplemental Material

**Supplementary Table 1.** Laboratory blood test parameters

| Blood parameter | Unit |
|---|---|
| Sodium | mmol/l |
| Potassium | mmol/l |
| Chloride | mmol/l |
| Total calcium | mmol/l |
| Corrected calcium | mmol/l |
| Total magnesium | mmol/l |
| Phosphate | mmol/l |
| Osmolality | mmol/Kg $H_2O$ |
| Calculated osmolality | mmol/Kg $H_2O$ |
| Osmolar gap | mmol/Kg $H_2O$ |

| | |
|---|---|
| Venous glucose | mmol/l |
| Creatinine (Jaffé) | µmol/l |
| Urea | mmol/l |
| Total cholesterol | mmol/l |
| HDL | mmol/l |
| Non-HDL | mmol/l |
| Triglycerides | mmol/l |
| Calculated LDL-cholesterol | mmol/l |
| Urate | µmol/l |
| Total bilirubin | µmol/l |
| Direct bilirubin | µmol/l |
| Iron | µmol/l |
| Albumin | g/l |
| CRP | mg/l |
| Transferrin | µmol/l |
| Transferrin saturation coef. | |
| Ferritin | µg/l |
| Hs-troponin T | ng/l |
| CK | U/l |
| CK-MB | U/l |

| | |
|---|---|
| LDH | U/l |
| AST (GOT) | U/l |
| ALT(GPT) | U/l |
| ϒ-GT | U/l |
| Pancreatic amylase | U/l |
| Lipase | U/l |
| Alkaline phosphatase | U/l |
| Measured bicarbonate | mmol/l |
| Anion gap | mmol/l |
| eGFR (CKD-EPI creatinine) | ml/min/1.73m$^2$ |
| NT-proBNP | ng/l |
| Haptoglobin | g/l |
| Procalcitonin | µg/l |
| Serum proteins | g/l |
| TSH | mUI/l |
| Total T4 | nmol/l |
| Free T4 | pmol/l |
| Total T3 | nmol/l |
| Free T3 | pmol/l |
| Reverse T3 | nmol/l |

| | |
|---|---|
| Prealbumin | g/l |
| Free carnitine | µmol/l |
| Total carnitine | µmol/l |
| Acyl-carnitine | µmol/l |
| Acyl-Carnitine/Free Carnitine | mol/mol |
| Acylcarnitine's profil | µmol/l |
| Carnitine (C0) | µmol/l |
| Acetyl carnitine (C2) | µmol/l |
| Propionyl carnitine (C3) | µmol/l |
| Malonyl carnitine (C3DC) | µmol/l |
| Methyl malonyl carnitine (C32MDC) | µmol/l |
| Butyryl carnitine (C4) | µmol/l |
| Hydroxy butyryl carnitine (C4OH) | µmol/l |
| Succinyl carnitine (C4DC) | µmol/l |
| 3-hydroxyisovalerylcarnitine/2-methyl-3-hydroxybutyrylcarnitine (C5OH) | µmol/l |
| Tiglylcarnitine/3-methylcrotonylcarnitine (C5:1) | µmol/l |
| Isovalerylcarnitine/2-methylbutyrylcarnitine (C5) | µmol/l |
| Glutaryl carnitine (C5DC) | µmol/l |
| Hexanoyl carnitine (C6) | µmol/l |
| 3-hydroxyhexanoycarnitine (C6OH) | µmol/l |

| | |
|---|---|
| Adipyl carnitine/3-methylglutarylcarnitine (C6DC) | µmol/l |
| Heptanoyl carnitine (C7) | µmol/l |
| Octenyl carnitine (C8:1) | µmol/l |
| Octanol carnitine (C8) | µmol/l |
| 3-hydroxyoctanylcarntine (C8OH) | µmol/l |
| Suberoylcarnitne (C8DC) | µmol/l |
| Decadienoylcarnitine (C10:2) | µmol/l |
| Decenylcarnitine (C10:1) | µmol/l |
| Decanoyl carnitine (C10) | µmol/l |
| 3-hydroxydecanoylcarnitine (C10OH) | µmol/l |
| Dodecenoylcarnitine (C12:1) | µmol/l |
| Dodecanoyl carnitine (C12) | µmol/l |
| 3-hydroxydodecanoylcarnitine (C12OH) | µmol/l |
| Tetradecadienoylcarnitine (C14:2) | µmol/l |
| Tetradecenoyl carnitine (C14:1) | µmol/l |
| Tetradecanoyl carnitine (C14) | µmol/l |
| 3-hydroxytetradecenoyalcarnitine (C14:1OH) | µmol/l |
| 3-hydroxytetradecanoylcarnitine (C14OH) | µmol/l |
| Palmitoleylcarnitine (C16:1) | µmol/l |
| Palmitoyl carnitine (C16) | µmol/l |

| | |
|---|---|
| 3-hydroxypalmitoleylcarnitine (C16:1OH) | µmol/l |
| 3-hydroxypalmitoylcarnitine (C16OH) | µmol/l |
| Linoleylcarnitine (C18:2) | µmol/l |
| Oleyl carnitine (C18:1) | µmol/l |
| Stearoyl carnitine (C18) | µmol/l |
| 3-hydroxylinoleylcarnitine (C18:2OH) | µmol/l |
| 3-hydroxyoleylcarnitine (C18:1OH) | µmol/l |
| 3-hydroxystearoylcarnitine (C18OH) | µmol/l |
| C3/C4 ratio | |
| C4/C2 ratio | |
| C5/C3 ratio | |
| C5DC/C3 ratio | |
| C8/C10 ratio | |
| C8/C10:1 ratio | |
| C14:1/C18:1 ratio | |
| C16OH/C16 ratio | |
| C0/(C16+C18) ratio | |
| C2/(C16+C18) ratio | |
| C14:1/C12:1 ratio | |
| (C16+C18:1)/C2 ratio | |

| | |
|---|---|
| Leukocytes | G/l |
| Erythrocytes | T/l |
| Haemoglobin | g/l |
| Haematocrit | % |
| MCV | fl |
| MCH | pg |
| MCHC | g/l |
| RDW | % |
| Platelets | G/l |
| Mean platelet volume | fl |
| Platelet distribution width | fl |
| Erythroblasts (automated) | /100 leuko. |
| Reticulocytes | °/oo |
| Absolute reticulocytes | G/l |
| Immature reticulocyte fraction | % |
| Immature reticulocytes | % |
| Haemoglobin concentration in reticulocytes | pg |

ALT: alanine aminotransferase; AST: aspartate aminotransferase; CK: creatine kinase; CK-MB: creatine kinase MB isoenzyme; CRP: c-reactive protein; eGFR: estimated glomerular filtration rate; HDL: high-density lipoprotein; Hs-troponin T: high-sensitivity troponin T; LDH: lactate dehydrogenase; LDL: low-density lipoprotein; MCH: mean corpuscular hemoglobin; MCHC: mean corpuscular hemoglobin concentration; MCV: mean corpuscular volume; MPV: mean platelet volume; NT-proBNP: N-terminal prohormone of b-type natriuretic peptide; PDW: platelet distribution width; RDW: red cell distribution width; TSH: thyroid-stimulating hormone; γ-GT: gamma-glutamyl transferase.

**Supplementary Table 2.** Echocardiographic parameters by imaging window

| Imaging window | Linear measurements | M-Mode/Areas/Volumes | Color Doppler | Spectral Doppler |
|---|---|---|---|---|
| Parasternal long axis | Septal thickness (mm)<br>Posterior wall thickness (mm)<br>LV diastolic diameter (mm)<br>LV systolic diameter (mm)<br>Left atrial diameter (mm)<br>LVOT diameter (mm)<br>Aortic root diameter (mm)<br>Sino-tubular junction (mm)<br>Ascending aorta (mm)<br>RVOT diameter (mm) | | Vena contracta (if AR or MR present) | |

| | | | | |
|---|---|---|---|---|
| Parasternal RV inflow view | | | Vena contracta (if TR present)<br>PISA radius (if TR present) | Tricuspid regurgitation (CwD) |
| Parasternal short axis (aortic) | Main pulmonary artery (mm)<br>RVOT diameter (mm) | Aortic valve planimetry | Vena contracta (if TR present)<br>PISA radius (if TR present) | Tricuspid regurgitation (CwD)<br>Pulmonary valve flow (CwD)<br>RVOT flow (PwD) |
| Parasternal short axis (mitral) | | Mitral valve planimetry | MR ellipticity | |
| Parasternal SAX (papillary) | | | | |
| Parasternal short axis (apex) | | | | |
| Apical 4-chambers | | Left atrial volume<br>Diastolic LV volume<br>Systolic LV volume | Vena contracta (if MR present)<br>PISA radius (if MR present) | Mitral inflow leaflet tip (PwD)<br>Mitral inflow annulus (PwD) |

| | | | | |
|---|---|---|---|---|
| | | | | Pulmonary vein flow (PwD)<br><br>Mitral inflow (CwD)<br><br>Mitral regurgitation (CwD)<br><br>Septal mitral annulus (TD)<br><br>Lateral mitral annulus (TD) |
| Apical 2-chambers | Descending aorta diam (mm) | Left atrial volume<br><br>Diastolic LV volume<br><br>Systolic LV volume | | |
| Apical 3-chambers | | | Vena contracta (if MR present)<br><br>PISA radius (if MR present) | |
| Apical 5-chambers | | | Vena contracta (if AR present)<br><br>PISA radius (if AR present) | Aortic outflow (CwD)<br><br>Aortic regurgitation flow (CwD) |

| | | | | |
|---|---|---|---|---|
| | | | | LVOT flow (PwD) |
| RV-modified 4-chambers view | RV basal diameter (mm)<br><br>RV mid diameter (mm)<br><br>RV length (mm) | Right atrial volume<br><br>Right atrial area<br><br>RV diastolic area<br><br>RV systolic area<br><br>TAPSE | Vena contracta (if TR present)<br><br>PISA radius (if TR present) | Tricuspid inflow (PwD)<br><br>Tricuspid inflow (CwD)<br><br>Tricuspid regurgitation (CwD)<br><br>Tricuspid annulus (TD) |
| Sub-costal long axis | | | | Tricuspid regurgitation (CwD) |
| Sub-costal short axis | Same as parasternal SAX | | | |
| Vena cava long axis view | Diameter exspiration (mm)<br><br>Diameter inspiration (mm)<br><br>Diameter snif (mm) | M-Mode IVC diameter | | Hepatic vein flow (PwD) |
| Abdominal aorta (SAX/LAX) | Diameter (mm) | | | Flow in AR present (PwD) |

| | | | | |
|---|---|---|---|---|
| Supra-sternal | Aortic arch diameter (mm) | | | Aortic arch flow (CwD)<br><br>Aortic arch flow (PwD) if AR present |
| Pedoff probe (suprasternal, apical/right parasternal | | | | Aortic outflow (CwD) if AS present |

AR: aortic regurgitation; AS: aortic stenosis; CwD: continuous wave doppler; IVC: inferior vena cava; LAX: long axis; LV: left ventricle; LVOT: left ventricular outflow tract; MR: mitral regurgitation; PwD: pulsed wave doppler; PISA: proximal isovelocity surface area; RA: right atrium; RV: right ventricle; RVOT: right ventricular outflow tract; SAX: short axis; TD: tissue doppler; TAPSE: tricuspid annular plane systolic excursion; TR: tricuspid regurgitation.

**Supplementary Table 3.** Measured and calculated MRI Parameters for each pulse sequence. Units are mentioned in parentheses when relevant, as is the subdivision. The subdivisions of the CMR parameters are: WH whole heart, 3P three planes, AHA16/AHA17 the 16 or 17 segments defined by the American Heart Association (i.e. excluding the apex or not), and S-D means quantified for both systole and diastole.

| Sequence | Images | Measurements |
|---|---|---|
| HASTE | Stack of 2D $T_2$-weighted kidney images | Kidney volume ($mm^3$), cortex volume, medulla volume, cortex diameter |
| BOLD | Single 2D R2*map | Cortex $T_2$* (ms), Medulla $T_2$* (ms) |
| Kidney PARMANav | Single 2D $T_1$&$T_2$ map | Cortex $T_1$ (ms), Medulla $T_1$ (ms), Cortex $T_2$ (ms), Medulla $T_2$ (ms) |
| Heart PARMANav | 4 2D $T_1$&$T_2$ maps, before and after GBCA injection | Myocardial $T_1$ (ms, WH/3P/AHA17), Myocardial $T_2$ (ms, WH/3P/AHA17), Myocardial ECV (ms, WH/3P/AHA17), Blood $T_1$ (ms) |
| 2D cine | Stack of 10-12 dynamic 2D heart images | Myocardial volume (mL, AHA17), LV volume (mL, S-D), RV volume (mL, S-D), LV EF (%), RV EF (%), LV stroke volume (mL), RV stroke volume (mL) |

| | | |
|---|---|---|
| qPerf | 3 dynamic GBCA perfusion images and 3 quantitative perfusion maps, under pharmacological stress and at rest | Stress perfusion (mL/min/g, WH/3P/AHA16), Rest perfusion (mL/min/g, WH/3P/AHA16), Myocardial perfusion reserve (-,WH/3P/AHA16) |
| 5D FRF | 5D (3D+respiratory&cardiac cycles) $T_1$-weighted whole-heart image | Myocardial volume (mL, AHA17), LV volume (mL, S-D), RV volume (mL, S-D), LV EF (%), RV EF (%), LV stroke volume (mL), RV stroke volume (mL) |
| 4D flow | 4D (3D spatial + cardiac cycle) flow in 3 directions | Asc. aorta peak flow (mL/s), Asc. aorta net flow volume (mL), Desc. aorta peak flow (mL/s), Desc. aorta net flow volume (mL), Main pulmonary artery peak velocity (cm/s), Main pulmonary artery net flow volume (mL), Mitral valve inflow peak velocity (cm/s), Mitral valve inflow net flow volume (mL), Mitral regurgitant fraction (%), Tricuspid valve inflow peak velocity (cm/s), Tricuspid valve inflow net flow volume (mL), Tricuspid |

| | | |
|---|---|---|
| | | regurgitant fraction (%), Left ventricle outflow net flow volume (mL), Right ventricle outflow net flow volume (mL) |

**Supplementary Table 4**. Collected clinical parameters.

| | |
|---|---|
| Demographic parameters | |
| Year of birth | |
| Sex | |
| Ethnicity | |
| Anthropometric parameters | |
| Weight | Kg |
| Height | Cm |
| Body mass index | $Kg/m^2$ |
| Consumption habits | |
| Smoking | Yes/No |
| If yes | |
| Years of smoking | Year |
| Number of cigarettes | Number/Day |
| Alcohol | Yes/No |
| If yes | |
| Years of alcohol use | Year |
| Number of standard drinks per occasion | |
| Drug | Yes/No |
| Medical history | |

| | |
|---|---|
| Cardiovascular | |
| Coronary disease | Yes/No |
| Myocardial infarction | Yes/No |
| Family history of coronary artery disease | Yes/No |
| Previous hospitalization for heart failure | Yes/No |
| Aortic stenosis | Yes/No |
| Aortic regurgitation | Yes/No |
| Mitral stenosis | Yes/No |
| Mitral regurgitation | Yes/No |
| Tricuspid regurgitation | Yes/No |
| Equipped with pacemaker or cardiac implant | Yes/No |
| Dyslipidaemia | Yes/No |
| Atrial fibrillation | Yes/No |
| Peripheral artery disease | Yes/No |
| Other | |
| Medication history | |
| Medication | Yes/No |
| If yes | |
| Name(s) of the medication(s) | |
| Frequency | |

| | |
|---|---|
| Dosage | |
| Indication | |
| Women only | |
| Ever used hormonal contraception | Yes/No |
| If yes | |
| Name of the contraception | |
| Dosage | |
| Route of administration | |
| Menopause | Yes/No |
| If yes | |
| Ever used hormone replacement therapy (HRT) | Yes/No |
| COMORBIDITIES | |
| Obesity | Yes/No |
| Diabetes mellitus | Yes/No |
| Hypertension | Yes/No |
| Renal dysfunction | Yes/No |
| Anemia | Yes/No |
| Iron deficiency | Yes/No |
| Sleep disorder | Yes/No |
| Chronic obstructive pulmonary disease (COPD) | Yes/No |

| VITAL SIGNS | |
|---|---|
| Temperature | ℃ |
| Heart rate | bpm |
| Blood pressure (systolic/diastolic) | mmHg |
| Respiratory rate | bpm |
